# Variant rs9644568 in the intergenic region downstream of the LPL gene is associated with high LDL cholesterol levels among Filipinos

**DOI:** 10.1101/2024.05.12.24307253

**Authors:** Eva Maria C. Cutiongco–de la Paz, Jose B. Nevado, Lourdes Ella G. Santos, Aimee Yvonne Criselle L. Aman, Jose Donato A. Magno, Paul Ferdinand M. Reganit, Richard Henry P. Tiongco, Deborah Ignacia D. Ona, Felix Eduardo R. Punzalan, Elmer Jasper B. Llanes, Jaime Alfonso M. Aherrera, Carla Mae O. Fernandez, Lauro L. Abrahan, Charlene F. Agustin, Adrian John P. Bejarin, Rody G. Sy

## Abstract

High blood level of low-density lipoprotein cholesterol (LDL-C) is a major risk factor for cardiovascular disease. Although genetic variants linked to high LDL-C have been studied in other populations, there have been no previous studies among Filipinos. This study aims to determine the association of candidate genetic variants to high LDL-C. We performed an age- and sex-matched case-control study that compared Filipino participants with high LDL-C levels (n=60) with controls (n=60). DNA was extracted from blood samples and genotyped for candidate SNPs using a customized microarray chip. Logistic regression analyses were used to determine the composite association of genetic and clinical variables to the condition. Of the initial eleven SNPs associated with high LDL-C in univariate analyses, only the variant rs9644568 in the intergenic region downstream of the *LPL* gene remained significantly associated with high LDL-C levels on multiple regression analysis and variable selection after adjustment for hypertension. The G allele was observed as the risk allele in a recessive model. The variant rs9644568-G in the *LPL* gene was associated with high blood LDL-C levels among Filipinos. In combination with hypertension, this genetic profile may identify individuals who are susceptible to develop high LDL-C in this population.

## Introduction

Low-density lipoprotein cholesterol (LDL-C) level is a risk factor and a traditional target for preventive cardiovascular disease (CVD) interventions. This association is enhanced in the presence of atherosclerosis [1], and the common view is that it is an active component of atherogenesis [2]. Several studies have since reported reduced adverse cardiovascular events after treatment with LDL-C-lowering drugs [2-4].

LDL-C levels may be affected by several factors, including genetics. A common familial form of LDL-C elevation, familial hypercholesterolemia (FH), has been well studied and three causative genes have already been identified, namely *low-density lipoprotein receptor (LDLR), apolipoprotein B (APOB), and proprotein convertase subtilisin/kexin type 9 (PCSK9)*. In addition, several studies on hereditary dyslipidemias have shown a variety of genes involved, such *as low density lipoprotein receptor adaptor protein 1 (LDLRAP1), ATP binding cassette subfamily G members 5 and 8 (ABCG5, ABCG8), angiopoietin like 3 (ANGPTL3), microsomal triglyceride transfer protein (MTTP), myosin regulatory light chain interacting protein (MYLIP), apolipoproteins A5, B, C2 and E (APOA5, APOB, APOC2, APOE), and lipoprotein lipase (LPL)*, which as expected, are involved in lipid metabolism, transport and processing [5,6].

Genotyping efforts in recent years attempted to find possible genetic susceptibility loci. For instance, genome-wide association studies (GWAS) have implicated *LDLR, APOE*, and *3-hydroxy-3-methylglutaryl-coenzyme A reductase* (*HMGCR*) as possible key regulators of lipid metabolism to high LDL-C [7-9]. Other genes that were considered include *fatty acid desaturase 1* (*FADS1*), *polypeptide N-acetylgalactosaminyltransferase 2* (*GALNT2*), and a*cetyl-coenzyme A carboxylase beta* (*ACACB*) [10-12].

However, findings from studies of genetic polymorphisms cannot be generalized across populations due to inherent inter-ethnic differences in genetic profiles. To illustrate, among the Taiwanese population, a *PCSK9* variant has been determined to have a causally significant association with high LDL-C levels; however, this association is not found among the Italian population [13,14]. In another study, a novel polymorphism of the *ICAM1* was reported to be associated with LDL-C in African Americans at conventional genome-wide significance, but not among Hispanics and East Asians [15]. Such discrepancies warrant verifying associations between genotypes and LDL-C levels in newly investigated populations. This could be due to independently evolved genetic differences with similar phenotypes due to metabolic proximity. Data from different ethnic populations will be able to contribute to a better understanding of diseases and can enhance customized clinical applications for individual populations.

Due to scarce data on genetic associations with LDL-C among Filipinos, this study aimed to identify candidate genetic markers associated with high LDL-C levels in this population. In addition to direct association, the variants were interpreted in the context of traditional risk factors to account for their significant, but potentially interdependent influences.

## Methodology

### Study design and ethics clearance

This is an age- and sex-matched case-control study performed from July 9, 2013 to March 31, 2017. This study complied to the Principles of the Declaration of Helsinki (2013) and all experimental protocols in this study were approved by the University of the Philippines Manila-Research Ethics Board (Study protocol code UPMREB-2012-0186-NIH). The investigators confirm that the methods complied with the relevant guidelines and regulations of UPMREB.

### Enrollment of participants and clinical data collection

Participants were enrolled from the Philippine General Hospital, different local communities, and clinics in Metro Manila. They were unrelated, aged 18 years old and above, of Filipino descent up to the 3^rd^ degree of consanguinity, and could provide independent consent. Cases were defined as (a) having at least one reading of serum LDL-C ≥160 mg/dl as defined by the NCEP ATP III guidelines; and (b) had not ever been on antilipidemics. Controls were participants with (a) at least one reading of serum LDL-C <160 mg/dl; and (b) without a previous diagnosis of dyslipidemia. Participants were excluded if they had end-stage renal disease, decompensated chronic liver disease, active malignancy, chronic lung disease, secondary hypertension, secondary dyslipidemia, or if pregnant. The demographic data and clinical characteristics were obtained from patient records and verbal interviews. Clinical data, including lipid profile and serum creatinine, were also recorded. Written informed consent forms duly approved by the UPMREB was obtained from potential participants prior to recruitment to the study.

### DNA extraction and quantification

DNA was extracted using the QiaAmp DNA Mini Kit following the spin protocol for blood buffy coat as specified in the manufacturer’s instruction manual. DNA was quantified using a spectrometer at 260nm and stored at -20°C until use. All DNA samples had A-260nm/A-280nm values above 1.80.

### Customized genotyping

A customized GoldenGate Genotyping (GGGT) beadchip (Illumina, Inc., San Diego, CA, USA) was designed in 2012 using candidate SNPs from both coding and noncoding regions, including intergenic and intronic SNPs, which have shown evidence of association with high LDL-C levels and lipid metabolism. These were selected after an extensive search in the following databases: Pharmacogenomics Knowledgebase database (PharmGKB), National Human Genome Research Institute Genome-Wide Association Study Catalog (NHGRI GWAS Catalog), PubMed, and selected international patent databases (e.g., Patentscope and Espacenet). The selected SNPs were submitted to Illumina, Inc. for scoring to determine the suitability of the SNPs to discriminate for microarray assay and to estimate their specificity.

Customized genotyping of candidate SNPs was performed using DNA microarray technology following the GoldenGate Genotyping (GGGT) protocol specified in the manufacturer’s manual. While the microarray platform is optimally designed to detect bi-allelic SNPs, the study included tri-or quad-allelic SNPs using the reference allele and the most common alternative in cases where the variants correlate with significant clinical outcomes. After microarray processing, the beadchip was imaged on the HiScan System, and data from these images were analyzed using GenomeStudio software.

## Data analysis

### Quality control

GenomeStudio 2.0 and PLINK version 2.05.10 were used for data cleanup using the following criteria: call rates > 95%, minor allele frequency (MAF) of > 0.01, genotype missingness of < 0.05, individual missingness < 0.05, and not having a Hardy-Weinberg equilibrium (HWE) of p < 0.001 in the control group.

### Statistical analysis

Chi-squared test and Fisher exact test, as applicable, were used for categorical data, and the student *t-test* was used for quantitative data were performed to assess for significant differences between alleles and genotypes (i.e., allelic and genotypic association tests, which include additive, dominant and recessive models). These models were identified based on the distribution of the case and control genotypes. Tests for allelic and genotypic associations were done by setting the cut-off at Bonferroni-corrected p < 0.05.

Upon determination of genotypic effects, the genotypes were recorded according to the most significant model, with imputations made for missing data. Univariate analysis was done to determine the association of SNPs with high LDL-C phenotype. A multiple logistic regression using stepwise backward elimination was performed with nominal p < 0.05 and clinical variable with p < 0.20.

## Results

After data quality control, the final set of samples for analysis included 120 participants consisting of 60 cases and 60 controls (Fig 1A). Using age- and sex-matched case-control design, the 120 participants were genotyped using 132 genomic variants.

**FIG 1.**
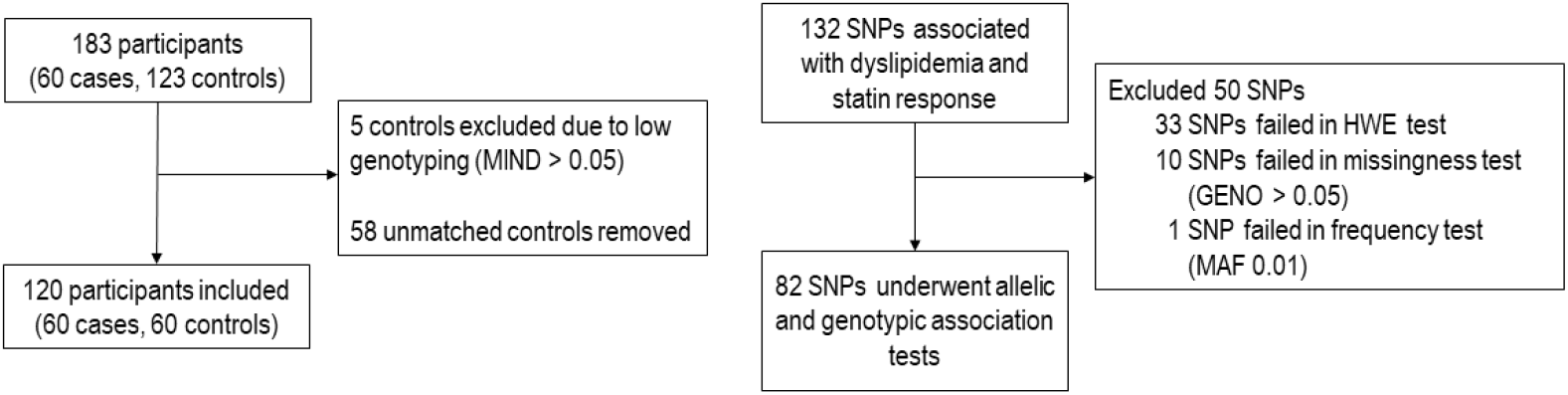
Schematic diagram of data processing and analysis for the high LDL-C group. A total of 183 participants (A) and 132 SNPs (B) were analyzed to determine the association of genetic variants with high LDL-C and lipid metabolism. Abbreviations: MIND – individual missingness; SNP – single nucleotide polymorphism; HWE – Hardy-Weinberg equilibrium; GENO – genotypic missingness

Of the 132 variants selected for the study, 50 variants were removed after quality control with PLINK: 33 SNPs failed the Hardy-Weinberg test (*p* > 0.001) among controls, ten SNPs had missing genotype data for more than 5% of the individuals, and one SNP was removed because the minor allele frequency was lower than 1% (Fig 1B). Eleven variants were significantly associated with high LDL-C after adjusting for multiple testing (i.e., Bonferroni-adjusted α = 0.05).

Table 1 summarizes the clinical characteristics of the included participants. Their crude odds ratios are in Table S1. Participants in the high LDL-C group tend to have more comorbidities that showed significance such as hypertension (*p* < 0.0001), BMI ≥ 25 kg/m^2^ (*p* = 0.056), and ischemic heart disease (*p* = 0.002). The cases also seemed to have slightly elevated creatinine levels.

**TABLE 1.**
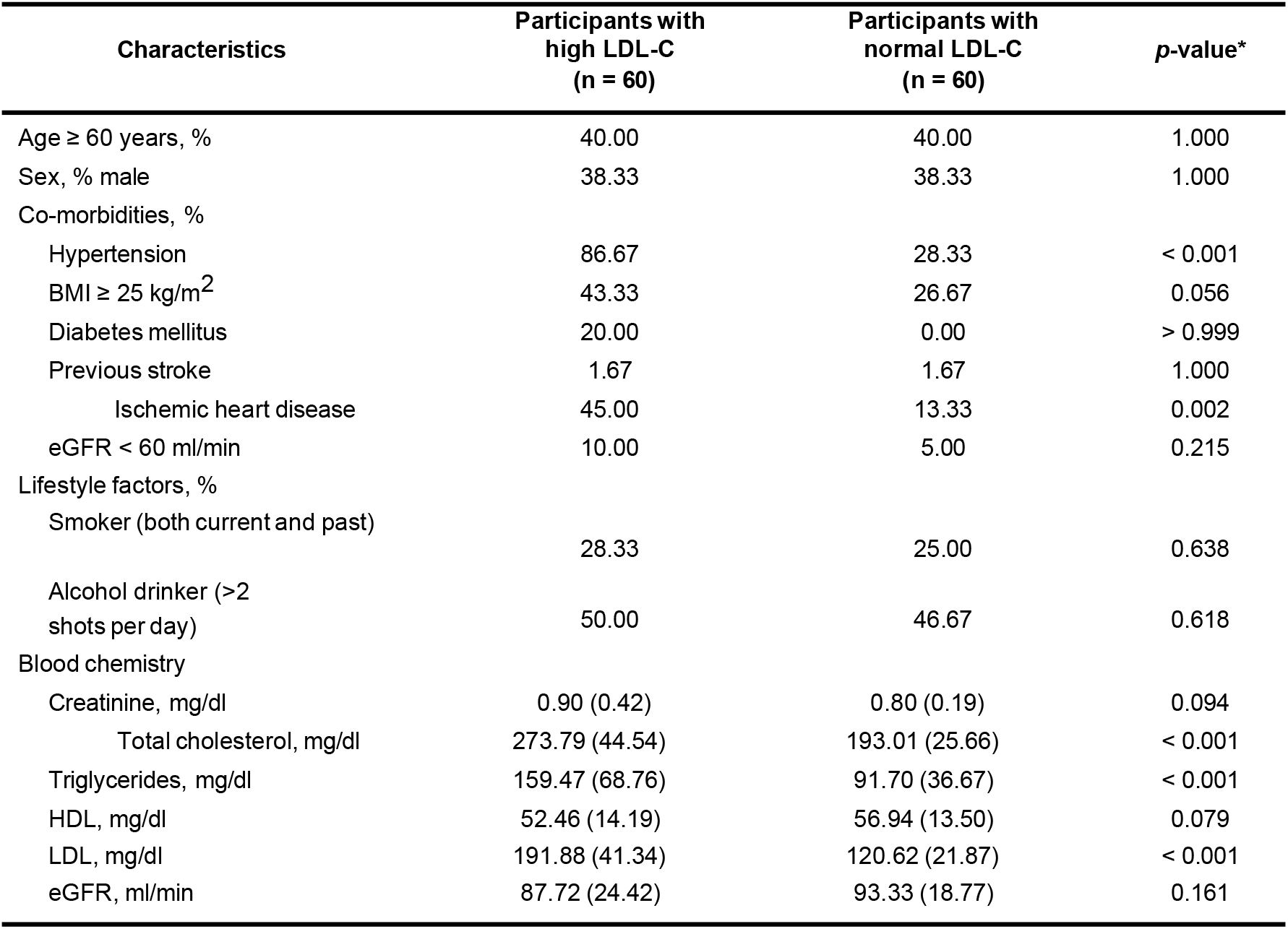
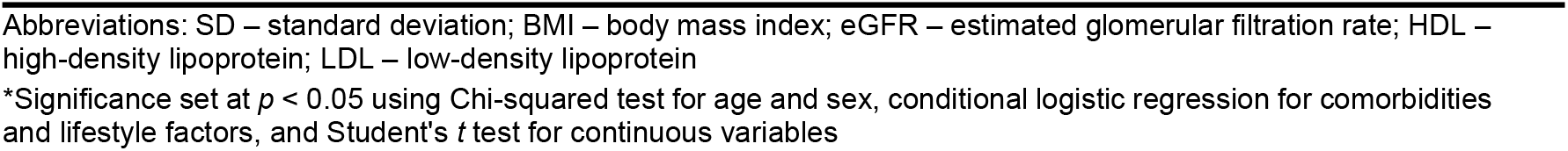
Clinical characteristics of participants included in the study.

| Characteristics | Participants with high LDL-C (n = 60) | Participants with normal LDL-C (n = 60) | p-value* |
| --- | --- | --- | --- |
| Age $\geq 60$ years, % | 40.00 | 40.00 | 1.000 |
| Sex, % male | 38.33 | 38.33 | 1.000 |
| Co-morbidities, % |  |  |  |
| Hypertension | 86.67 | 28.33 | < 0.001 |
| BMI $\geq 25$ kg/m<sup>2</sup> | 43.33 | 26.67 | 0.056 |
| Diabetes mellitus | 20.00 | 0.00 | > 0.999 |
| Previous stroke | 1.67 | 1.67 | 1.000 |
| Ischemic heart disease | 45.00 | 13.33 | 0.002 |
| eGFR < 60 ml/min | 10.00 | 5.00 | 0.215 |
| Lifestyle factors, % |  |  |  |
| Smoker (both current and past) | 28.33 | 25.00 | 0.638 |
| Alcohol drinker (>2 shots per day) | 50.00 | 46.67 | 0.618 |
| Blood chemistry |  |  |  |
| Creatinine, mg/dl | 0.90 (0.42) | 0.80 (0.19) | 0.094 |
| Total cholesterol, mg/dl | 273.79 (44.54) | 193.01 (25.66) | < 0.001 |
| Triglycerides, mg/dl | 159.47 (68.76) | 91.70 (36.67) | < 0.001 |
| HDL, mg/dl | 52.46 (14.19) | 56.94 (13.50) | 0.079 |
| LDL, mg/dl | 191.88 (41.34) | 120.62 (21.87) | < 0.001 |
| eGFR, ml/min | 87.72 (24.42) | 93.33 (18.77) | 0.161 |

Abbreviations: SD – standard deviation; BMI – body mass index; eGFR – estimated glomerular filtration rate; HDL – high-density lipoprotein; LDL – low-density lipoprotein
\*Significance set at $p < 0.05$ using Chi-squared test for age and sex, conditional logistic regression for comorbidities and lifestyle factors, and Student's $t$ test for continuous variables

There were 11 variants found to have a statistically significant association with LDL-C levels on the genotypic association test after adjusting for multiple testing (Bonferroni-adjusted *α* = 0.00061) (Table S2). These variants underwent logistic regression analysis for the computation of their OR and to determine which will retain significance upon adjusting for other variables (Table 2). One SNP had clinically significant odds ratios (OR ≥ 2.5) and was statistically significant (*p*<.001) on simple logistic regression analysis: rs12279250 in NELL1. This SNP seemed to display a dominant genotypic effect.

**TABLE 2.**
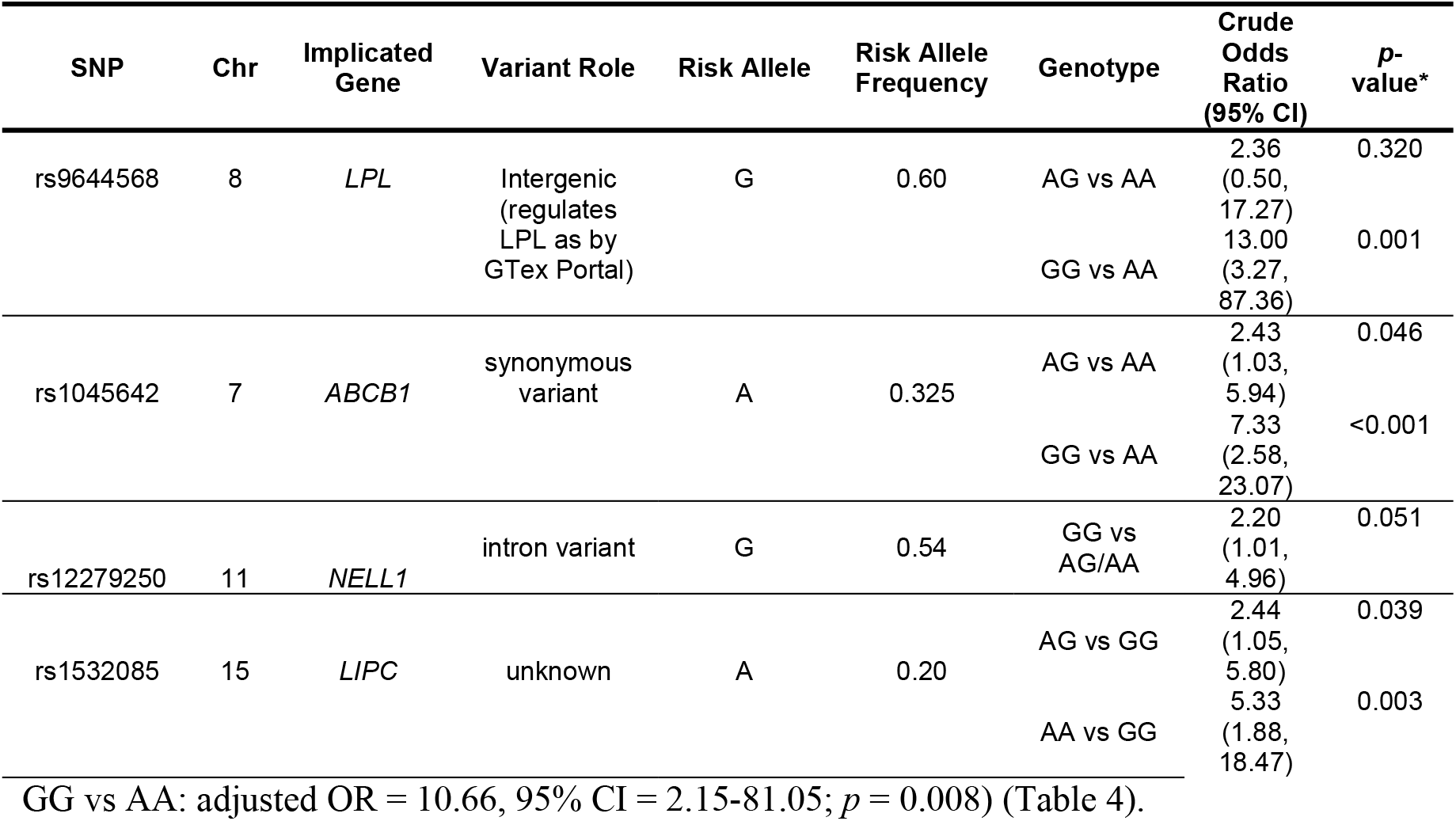
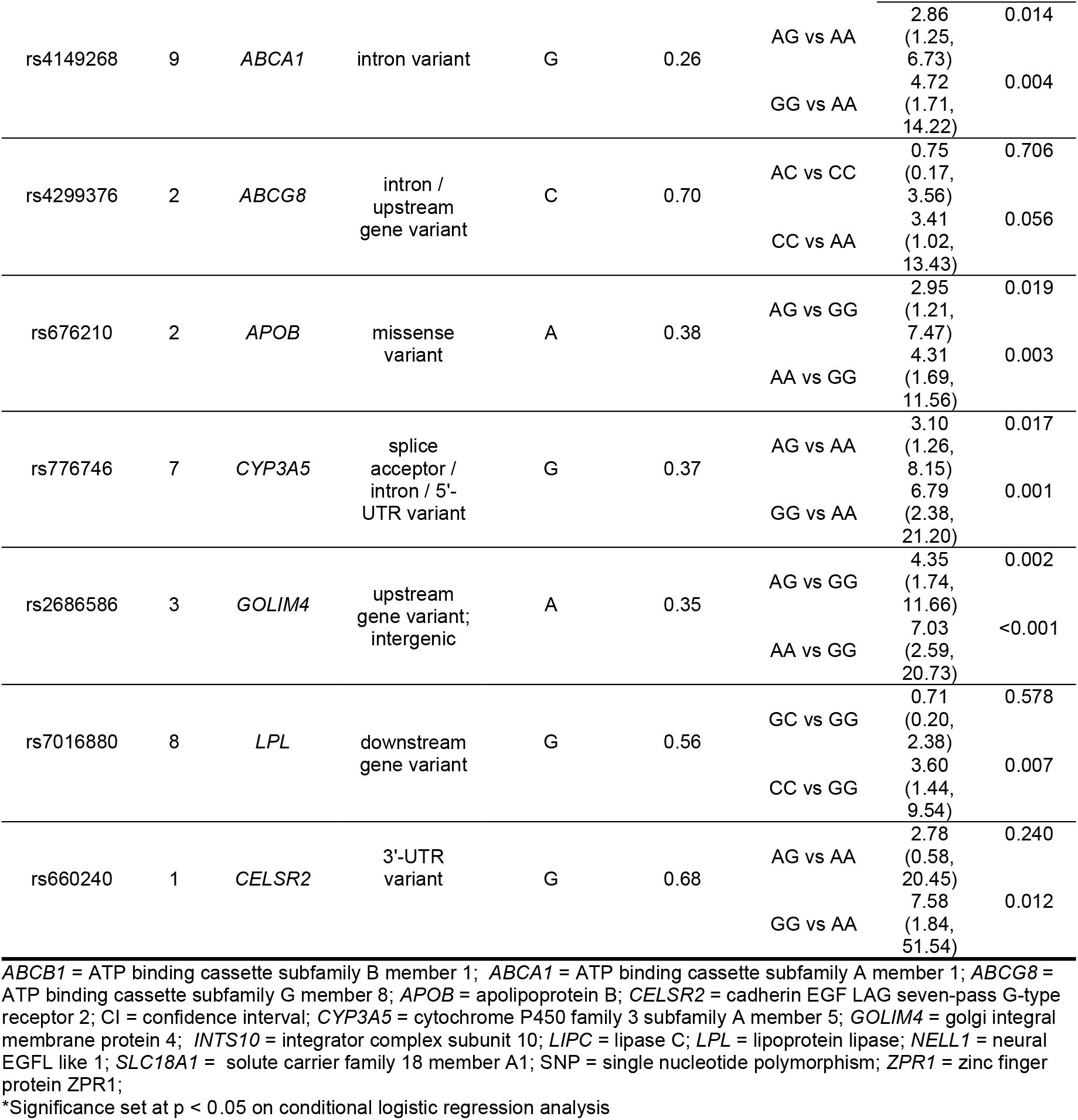
Univariate logistic regression analysis of the significant variants.

| SNP | Chr | Implicated Gene | Variant Role | Risk Allele | Risk Allele Frequency | Genotype | Crude Odds Ratio (95% CI) | $p$-value* |
| --- | --- | --- | --- | --- | --- | --- | --- | --- |
| rs9644568 | 8 | LPL | Intergenic (regulates LPL as by GTex Portal) | G | 0.60 | AG vs AA | 2.36 (0.50, 17.27) | 0.320 |
|  |  |  |  |  |  | GG vs AA | 13.00 (3.27, 87.36) | 0.001 |
| rs1045642 | 7 | ABCB1 | synonymous variant | A | 0.325 | AG vs AA | 2.43 (1.03, 5.94) | 0.046 |
|  |  |  |  |  |  | GG vs AA | 7.33 (2.58, 23.07) | <0.001 |
| rs12279250 | 11 | NELL1 | intron variant | G | 0.54 | GG vs AG/AA | 2.20 (1.01, 4.96) | 0.051 |
| rs1532085 | 15 | LIPC | unknown | A | 0.20 | AG vs GG | 2.44 (1.05, 5.80) | 0.039 |
|  |  |  |  |  |  | AA vs GG | 5.33 (1.88, 18.47) | 0.003 |
| GG vs AA: adjusted OR = 10.66, 95% CI = 2.15-81.05; $p = 0.008$ ) (Table 4). |  |  |  |  |  |  |  |  |
| rs4149268 | 9 | ABCA1 | intron variant | G | 0.26 | AG vs AA | 2.86<br>(1.25, 6.73) | 0.014 |
|  |  |  |  |  |  | GG vs AA | 4.72<br>(1.71, 14.22) | 0.004 |
| rs4299376 | 2 | ABCG8 | intron /<br>upstream<br>gene variant | C | 0.70 | AC vs CC | 0.75<br>(0.17, 3.56) | 0.706 |
|  |  |  |  |  |  | CC vs AA | 3.41<br>(1.02, 13.43) | 0.056 |
| rs676210 | 2 | APOB | missense<br>variant | A | 0.38 | AG vs GG | 2.95<br>(1.21, 7.47) | 0.019 |
|  |  |  |  |  |  | AA vs GG | 4.31<br>(1.69, 11.56) | 0.003 |
| rs776746 | 7 | CYP3A5 | splice<br>acceptor /<br>intron / 5'-<br>UTR variant | G | 0.37 | AG vs AA | 3.10<br>(1.26, 8.15) | 0.017 |
|  |  |  |  |  |  | GG vs AA | 6.79<br>(2.38, 21.20) | 0.001 |
| rs2686586 | 3 | GOLIM4 | upstream<br>gene variant;<br>intergenic | A | 0.35 | AG vs GG | 4.35<br>(1.74, 11.66) | 0.002 |
|  |  |  |  |  |  | AA vs GG | 7.03<br>(2.59, 20.73) | <0.001 |
| rs7016880 | 8 | LPL | downstream<br>gene variant | G | 0.56 | GC vs GG | 0.71<br>(0.20, 2.38) | 0.578 |
|  |  |  |  |  |  | CC vs GG | 3.60<br>(1.44, 9.54) | 0.007 |
| rs660240 | 1 | CELSR2 | 3'-UTR<br>variant | G | 0.68 | AG vs AA | 2.78<br>(0.58, 20.45) | 0.240 |
|  |  |  |  |  |  | GG vs AA | 7.58<br>(1.84, 51.54) | 0.012 |

*ABCB1* = ATP binding cassette subfamily B member 1; *ABCA1* = ATP binding cassette subfamily A member 1; *ABCG8* = ATP binding cassette subfamily G member 8; *APOB* = apolipoprotein B; *CELSR2* = cadherin EGF LAG seven-pass G-type receptor 2; CI = confidence interval; *CYP3A5* = cytochrome P450 family 3 subfamily A member 5; *GOLIM4* = golgi integral membrane protein 4; *INTS10* = integrator complex subunit 10; *LIPC* = lipase C; *LPL* = lipoprotein lipase; *NELL1* = neural EGFL like 1; *SLC18A1* = solute carrier family 18 member A1; SNP = single nucleotide polymorphism; *ZPR1* = zinc finger protein ZPR1;
\*Significance set at $p < 0.05$ on conditional logistic regression analysis

Clinical variables with a significant difference between the two groups, *p* < 0.20: hypertension, BMI ≥ 25 kg/m^2^, and ischemic heart disease were included together with the 11 variants with nominal *p <* 0.05 in the full model for the multiple regression analysis (Table 3). On variable selection using the stepwise algorithm (*p <* 0.05), only the presence of hypertension and rs9644568 retained their significant association with high LDL-C levels under an additive model with allele G as the risk allele (AG vs. GG: adjusted OR = 2.44, 95% CI = 0.41-20.60; *p* = 0.355;

**TABLE 3.**
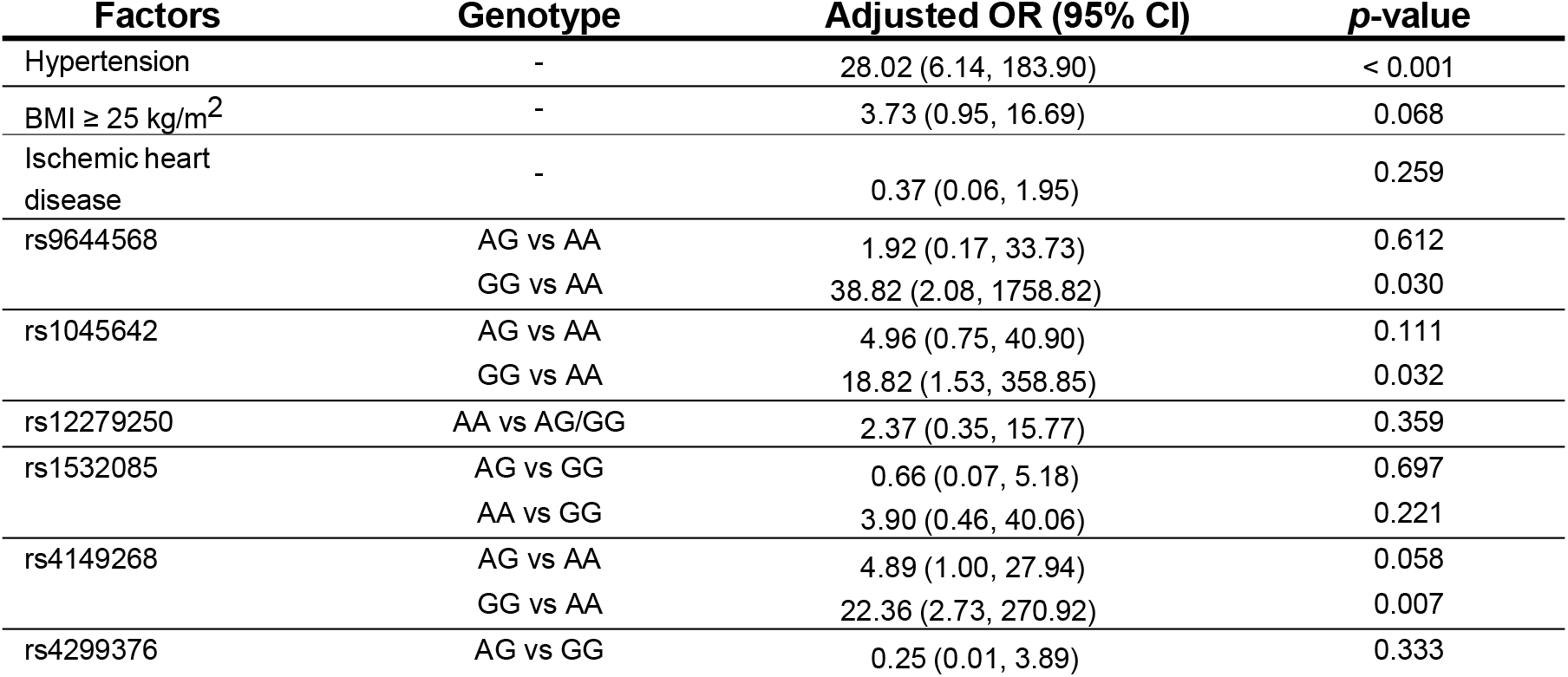
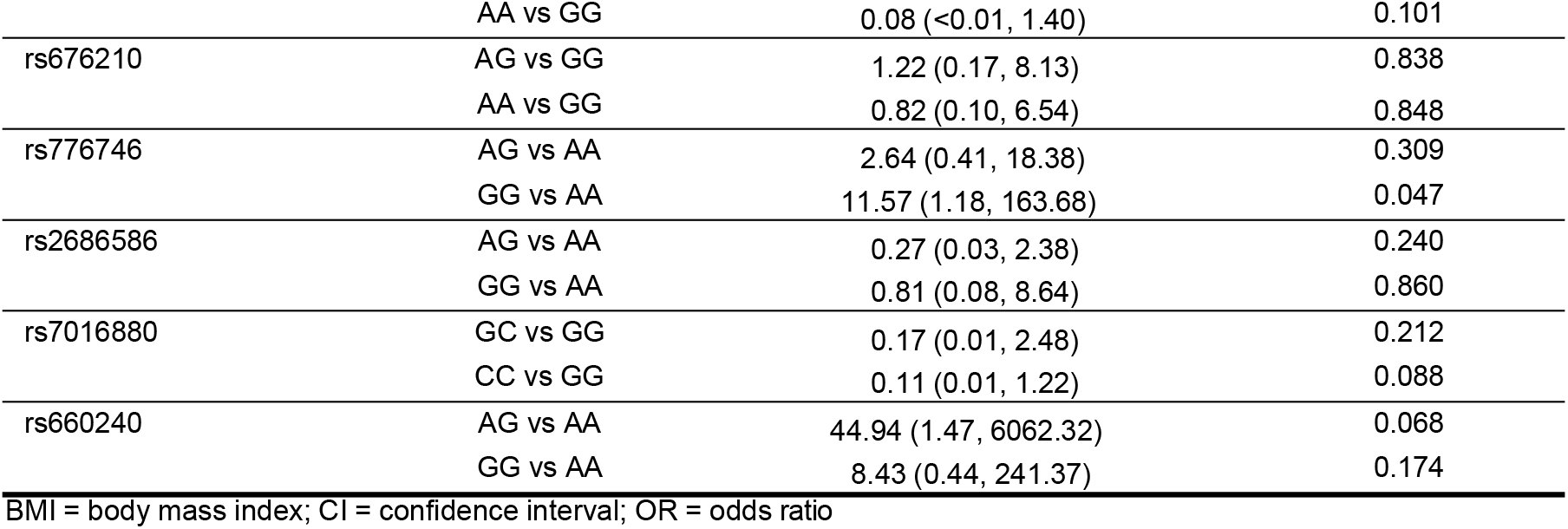
Genetic variants and clinical factors in the multiple regression full model.

**TABLE 4.** Genetic variants and clinical factors remaining after variable selection.

| Variable | Genotype | Adjusted OR (95% CI) | p-value |
| --- | --- | --- | --- |
| rs9644568 | AG vs AA | 2.44 (0.41, 20.60) | 0.355 |
|  | GG vs AA | 10.66 (2.15, 81.05) | 0.008 |
| Hypertension | - | 14.23 (5.53, 40.81) | < 0.001 |

\*Significance set at $p < 0.05$ ; Abbr: CI = confidence interval; OR = odds ratio

## Discussion

Genetic determinants of high LDL-C may be helpful in the understanding and optimum management of cardiovascular risk. As associated variations often differ across populations, especially for complex conditions, studies on Filipinos must be undertaken considering that cardiovascular disease remains as the top cause of mortality in the country. This study analyzes candidate genetic variants of high LDL-C among Filipino patients. In addition, a regression model of candidate genetic variants incorporating traditional clinical risk factors was generated to assess their relative effects on LDL-C profiles.

Univariate analyses found 11 variants associated with LDL-C levels. The reported genes include *lipoprotein lipase* (*LPL*), *ATP binding cassette subfamily B member* (*ABCB1*), *lipase C* (*LIPC*), *ATP binding cassette subfamily A member* (*ABCA1*), *ATP binding cassette subfamily G member 8* (*ABCG8*), *apolipoprotein B* (*APOB*), and *cadherin EGF LAG seven-pass G-type receptor 2* (*CELSR2*). Expectedly, these are known genes whose roles in LDL-C metabolism are well established — from lipid transport, cellular metabolism and homeostasis, cellular translocation, and storage [16-21].

A major finding in this study is the variant rs9644568 near the *LPL* gene. Logistic regression with adjustment for hypertension shows a significant association of rs9644568 to high LDL-C levels after adjusting for hypertension among Filipinos. The G allele of rs9644568 confers a 2.44-fold increased risk for high LDL-C in heterozygous individuals and a 10.66-fold increased risk in homozygous individuals. This suggests a possible additive or a multiplicative model of transmission. This is similar to the findings of a genome-wide association study among Europeans, which consistently demonstrated the significant association of G allele of rs4977574 with high LDL-C levels and an increased risk of myocardial infarction (MI) [22]. In contrast, the minor G allele protects LDL-C in another study that included Hispanics, East Asians and also Europeans [23]. A similar study found the minor G allele to be associated with decreased risk for coronary artery disease and peripheral artery disease in a Chinese population with diabetes [24].

The SNP rs9644568 is located on chromosome 8p21.3 in an intergenic region downstream of the gene *LPL* (UCSC Genome Browser, http://genome.ucsc.edu. Accessed October 10, 2023). Although the variant is >100 kb far from the gene LPL, expression from several tissues suggests that it regulates the gene (GTEx Portal. https://gtexportal.org. Accessed October 10, 2023).

This gene is predominantly expressed in the heart, skeletal muscles, and adipose tissues^25^. *LPL* encodes a rate-limiting enzyme called lipoprotein lipase, which plays an important role in lipid metabolism, as well as in lipid intake and clearance. Lipoprotein lipase aids in the breakdown of the circulating triglyceride-rich lipoproteins in the bloodstream and the uptake of free fatty acids into peripheral organs for energy storage and consumption [26-30]. Alterations in LPL activity could result in dyslipidemia, characterized by increased plasma LDL-C, TG, and reduced plasma HDL-C levels [31]. This association of the variant is interesting, as prior data, to our knowledge, do not indicate rs9644568 as a risk marker for high LDL levels or as having the highest odds ratio in any non-Filipino population. This can imply that the variant is a unique risk marker for Filipinos, independent of other components of metabolic syndrome, i.e., hypertension and diabetes mellitus.

To support the significance of *LPL* as a candidate locus, the study also indicated that another downstream *LPL* variant, rs7016880, was associated with LDL-C. Interestingly, rs7016880 shares many characteristics with rs9644568 – being located as a downstream non-coding regulator, having a possible multiplicative model, with a high odds ratio. This may provide greater confidence for the veracity of the findings in this study.

As for the other variants, the adjustments due to hypertension and the rs9644568 seem to mask their statistical significance. This has several implications. First, rs9644568 is rarely reported as a marker for LDL-C in the literature, but the statistical significance of rs9644568 stood out prominently in the current study, thus implying that the SNP is unique among the Filipino population. Second, hypertension might have masked the statistical significance of some of the candidate genes in the metabolic syndrome context. This is unsurprising as some variants have been previously associated with high blood pressure. To illustrate, *ABCBI* was reported to be associated with hypertension through dietary salt transport and retention [32]. In a study among Han Chinese, variants of *ABCA1*, rs2472510 and rs2515614, were associated with hypertension, with decreased expression of *ABCA1* linked with hypertension risk [33]. On the other hand, meta-analysis of 2799 cases and 6794 controls showed an association of *CYP3A5* rs776746 with hypertension [34]. A later study on a small sample of Han Chinese also reported a significant association of *CYP3A5* rs776746 with the elevated risk of hypertension [35].

Several variants are mostly known genes whose roles in LDL-C metabolism are well established [16-21]. The G allele of rs1532085 in *LIPC* with a protective effect for LDL-C levels in Chinese populations [36], is similar to the findings in this study. In a large GWAS study involving >100,000 Caucasians, rs4299376 in *ABCG8* was found to be one of the strongly associated genes with LDL-C [38]. The variant rs676210 in *APOB* has been identified as a susceptibility locus for very low-density lipoprotein levels in the Women’s Genome Health Study (WGHS) involving 17,296 women of European ancestry [38]. This association was further reported in the Chinese Yugur population^39^. A GWAS study of 11,685 showed a significant association of *CELSR2* rs660240 to LDL-C levels where the G allele is associated with risk and this was confirmed in a meta-analysis of Below et al. (2016) [40,41].

However, some genes can be considered as attractive at best at this point. One intriguing finding is the direct association of ABCB1 with LDL-C. There is scarce data on this association, as most studies focused on its association with response to statins [42,43]. The variant ABCB1 rs1045642 is by itself associated with statin response [44]. The variant rs12279250 in NELL1, a growth factor of still uncertain significance, has been associated with increased triglyceride levels [45,46], which may lead to a subsequent increase in LDL-C levels. Despite the study identifying rs4149268 in ABCA1 as associated with LDL-C, most studies reported an association with HDL-C levels instead of LDL-C [47]. This may need to be verified, but it is noteworthy to investigate if such an association remains among Filipinos. As for rs776746 in CYP3A5, there is little study on the direct association with LDL-C levels. This could be due to the assumption that cytochrome P proteins are mainly associated with drugs. In fact, most studies focused on response to cholesterol-lowering drugs [48,49]. As the present study focused on participants without drug intake, a possible role for CYP3A5 on LDL-C level could be plausible. Similarly, there is minimal information on rs2686586. Even its association with the gene GOLIM4 is questionable, as no evidence of regulation nor proximity (>50kb) is noted in the literature. Nonetheless, the fact that it is significantly associated with LDL-C needs to be verified.

In this study, clinical variables that show a significant association with high LDL-C phenotype are hypertension, BMI ≥ 25 kg/m2, and ischemic heart disease (IHD). Several studies consistently show strong correlations between these comorbidities and LDL-C levels. The link between LDL-C and hypertension was elucidated by the group of Laaksonen et al. (2008). They reported that elevated serum levels of LDL-C increase the risk of hypertension in Finnish men, which explained the strong association of dyslipidemia with incident hypertension [50]. Meanwhile, a previous study performed in the US population suggested a linear association of LDL-C concentrations and BMI [51]. The association between high LDL-C levels and high IHD risk is further substantiated by the study of Breuer (2005), focusing on improving cardiovascular disease outcomes by lowering LDL-C levels [52]. Moreover, dyslipidemia and hypertension are known as major risk factors for cardiovascular diseases; with these two comorbidities reported to be frequently diagnosed in patients with ischemic heart disease [53].

This study presents an opportunity to identify individuals at risk for developing high LDL-C levels by looking at their genetic profiles to complement clinical correlates. Estimated associations between variant rs9644568 and hypertension were very high, but the confidence intervals around them were too wide to rule out the possibility of substantial associations. Further studies with larger sample sizes are required to validate the conclusions of this study.

## Conclusion

Genetic determinants largely influence the development of high LDL-C levels. This study found an association of variant rs9644568 in the intergenic region downstream of the *LPL* gene, with high LDL-C levels among Filipinos. Adding a genetic component may improve screening and assessment of patients at risk for developing high LDL-C levels.

## Data Availability

All relevant data are within the manuscript and its Supporting Information files.

## Acknowledgements

The authors would like to express their sincere gratitude to the administrative staff and researchers of the Microarray Laboratory of the Institute of Human Genetics, National Institutes of Health-University of the Philippines Manila, the Philippine Genome Center, and the clinical fellows of the Department of Medicine for actively being involved in the project. We are extending our gratitude to our sponsor, the Philippine Council for Health Research and Development of the Department of Science and Technology (PCHRD-DOST)

## Author Contributions

**EMCutiongco-De La Paz**, conceptualized the study and designed the methodology, acquired funding and led the project administration and supervision, conducted the research and investigation process, performed the data curation and analysis, interpreted the data, wrote the original draft, reviewed and edited the manuscript for intellectual content; **JBNevado** conceptualized the study and designed the methodology, acquired funding and led the project administration and supervision, conducted the research and investigation process, performed the data curation and analysis, interpreted the data, wrote the original draft, reviewed and edited the manuscript for intellectual content; **LEGSantos**, conceptualized the study and designed the methodology, conducted the research and investigation process, interpreted the data; **AYCLAman**, acquired funding and led the project administration and supervision, conducted the research and investigation process, performed the data curation and analysis, interpreted the data, wrote the original draft; **JDAMagno**, conceptualized the study and designed the methodology, conducted the research and investigation process, interpreted the data; **PMMReganit**, conceptualized the study and designed the methodology, conducted the research and investigation process, interpreted the data; **RHPTiongco**, conceptualized the study and designed the methodology, conducted the research and investigation process, interpreted the data; **DIDOna**, conceptualized the study and designed the methodology, conducted the research and investigation process, interpreted the data; **FERPunzalan**, conceptualized the study and designed the methodology, conducted the research and investigation process, interpreted the data; **EJBLlanes**, conceptualized the study and designed the methodology, conducted the research and investigation process, interpreted the data; **JAMAherrera**, conducted the research and investigation process, interpreted the data; **CMOFernandez**, performed the data curation and analysis, interpreted the data, reviewed and edited the manuscript for intellectual content; **LLAbrahan**, conducted the research and investigation process, interpreted the data; **CFAgustin**, conducted the research and investigation process, interpreted the data; **AJPBejarin**, conducted the research and investigation process, performed the data curation and analysis, interpreted the data, wrote the original draft; **RGSy**, conceptualized the study and designed the methodology, acquired funding and led the project administration and supervision, conducted the research and investigation process, interpreted the data, reviewed and edited the manuscript for intellectual content

### Data Availability Statement

The datasets used and/or analyzed during the current study are available from the corresponding author on reasonable request.

### Funding Source

The present study was sponsored by a grant-in-aid from the PCHRD-DOST

